# The transition from primary colorectal cancer to isolated peritoneal malignancy is associated with a hypermutant, hypermethylated state

**DOI:** 10.1101/2020.02.24.20027318

**Authors:** Sally Hallam, Joanne Stockton, Claire Bryer, Celina Whalley, Valerie Pestinger, Haney Youssef, Andrew D Beggs

## Abstract

Coloretcal Peritoneal metastases (CPM) develop in 15% of colorectal cancers. Cytoreductive surgery and heated intraperitoneal chemotherapy (CRS & HIPEC) is the current standard of care in selected patients with limited resectable CPM. Despite selection using known prognostic factors survival is varied and morbidity and mortality are relatively high. There is a need to improve patient selection and a paucity of research concerning the biology of isolated CPM. We aimed to determine the biology associated with transition from primary CRC to CPM and poor prognosis CPM, to identify those suitable for treatment with CRS & HIPEC and to identify targets for existing repurposed or novel treatment strategies. A cohort of patients with CPM treated with CRS & HIPEC was recruited and divided according to prognosis. Molecular profiling of the transcriptome, epigenome and genome of CPM and matched primary CRC was performed.

CPM were characterised by frequent Wnt/ β catenin negative regulator mutations, mismatch repair mutations and resulting high tumour mutational burden and dysregulation of methylation suggested by frequent TET2 mutations and mutations suggesting an immune evasive phenotype. Several novel therapies could be targeted to these frequent mutations including porcupine inhibitors, immune checkpoint inhibitors and methylation inhibitors. Here we show the molecular features associated with CPM development and with poor prognosis. Potential applications include improving patient selection for treatment and in the development of novel and personalised treatments.

**NOVELTY AND IMPACT:** Colorectal peritoneal metastasis (CPM) are associated with limited and variable survival despite patient selection using known prognostic factors and optimal currently available treatments. There is a paucity of research concerning the biology of CPM. This study describes the biological landscape of CPM and the molecular features associated with CPM development, conferring poor prognosis and has identified that the majority of CPM develop a hypermutant phenotype that may be suitable for treatment with anti-PD1/CTLA4 immunotherapy.

## INTRODUCTION

Little is known about the biology of isolated colorectal peritoneal metastasis (CPM), which although a relatively rare phenomenon is one with a high mortality rate (1). Understanding tumour biology may identify which patients with primary colorectal cancer (CRC) are at risk of developing CPM, and which are suitable for treatment with cytoreductive surgery and heated intra-peritoneal chemotherapy (CRS & HIPEC). CRS & HIPEC (usually using an agent such as mitomycin C)aims to achieve macroscopic tumour resection with multiple visceral and peritoneal resections and ablation of microscopic disease. Five-year survival however varies widely, and morbidity and mortality are relatively high (1). There is a need therefore to improve patient selection, allowing alternative existing or novel treatment strategies to be used for patients unlikely to respond.

Primary CRC research has identified biological drivers of metastasis and markers of response to specific treatments, for example KRAS mutation in selection for anti-EGFR mAb therapy (2). Gene expression signatures can be used to predict oncological outcomes and treatment response, for example the CMS & CRIS classifiers of colorectal cancer biology (3, 4). Gene expression profiling in primary CRC has identified signatures associated with the development of metastasis (5). One small study combining a small number of CPM with a larger cohort of appendix adenocarcinoma identified a signature predictive of reduced overall survival (OS) following CRS & HIPEC; these are however two biologically distinct tumours, appendix having significantly improved prognosis (2).

The dysregulation of methylation is a key step in tumorigenesis CpG island promoter methylation (CIMP) appears to be stable between matched primary CRC and hepatic metastasis suggesting an epigenetic methylation programme is established prior to the development of metastasis (6, 7) (8). The hypermethylation of KRAS, Wnt modulators, tumour suppressor genes, CIMP and hypomethylation of the oncogenes are associated with an unfavourable response to chemotherapy and anti-EGFR antibodies as well as tumour recurrence and reduced OS in primary and metastatic CRC (9-14).Chromosomal instability is ubiquitous in cancer, increased copy number alteration, indicative of chromosomal instability is found in metastatic CRC (15, 16). Lopez-Garcia et al (17) demonstrated that the evolution of chromosomal instability is depending on cellular tolerance, either via dysregulation of TP53 or via alternate escape mechanisms such as dysfunction of BCL9L regulated caspase signalling.

CRC metastatic drivers are less clearly defined. Some studies found mutations exclusive to metastatic sites (18, 19), others found similar patterns of mutation between primary and metastasis (20). Studies have examined the somatic mutations in CPM and their prognostic implications. These studies are limited to individual or small panels of mutations routinely tested for in clinical practice with limited evidence to suggest which genes should be included in panel sequencing in CPM. Schneider et al examined the *KRAS* and *BRAF* mutation status of patients with CPM who underwent CRS & HIPEC (21). They found mutations of *RAS/RAF* were associated with reduced OS independent of the use of targeted anti-EGFR treatment (21). Sasaki et al examined the *KRAS, BRAF* and *PIKCA* mutation status of patients with metastatic CRC, with or without CPM (22). They found the incidence of *BRAF* mutation was significantly associated with the presence of CPM but not with prognosis (22).

The landscape of metastatic colorectal cancer was studied by the MSK-IMPACT (23) group which undertook panel based sequencing of 1,134 metastatic colorectal cancers. Of these 19 patients were defined as “peritoneal” malignancy which defined a wide range of anatomical sites (liver, ovary, abdomen) rather than pure peritoneal disease. These tumours were also not studied with matched primary tumour of origin, nor did they undergo comprehensive genomic characterisation, as early versions of the MSK panel were insufficient in size to call tumour mutational burden.

There is a need to improve the outcomes for patients with CPM and significant variation in survival despite patient selection for treatment using known prognostic factors. There is a paucity of knowledge concerning CPM tumour biology. Understanding tumour biology will identify patients with primary CRC at risk of developing CPM, those suitable for treatment with CRS & HIPEC or alternative existing and novel treatment strategies. This study aims to determine the landscape of gene expression, methylation, and somatic mutation profile associated with the transition from primary CRC to isolated CPM and determine the association between these and prognosis following CRS & HIPEC in order to identify therapeutic targets.

## MATERIALS AND METHODS

### Patient cohorts

This study obtained ethical approval from the North West Haydock Research Ethics Committee, (15/NW/0079), project ID (17/283). Participants gave informed consent. Consecutive retrospective patients were recruited from an internally held database of all patients undergoing CRS & HIPEC at Good Hope hospital from 2011–2017. Patients with CPM (adenocarcinoma), no extra-abdominal metastasis, a CC0 resection and a PCI of < 12 were eligible for inclusion. Patients were divided into two prognostic groups. With palliative chemotherapy DFS is 11-13 months and therefore patients, post-treatment (CRS & HIPEC) with disease free survival (DFS) <12 months were defined as “poor prognosis” (24). Patients undergoing therapy with DFS > 12 months were defined as “good prognosis”. Demographic, tumour and treatment details were compared between the prognostic cohorts. For continuous variables, the students T-test was applied to normally distributed data and Mann Whitney-U to non-normally distributed data. Categorical variables were compared with the Chi-squared test or Fishers exact test. A p-value of < 0.5 was considered statistically significant. DFS survival between the good and poor prognosis cohorts was compared using the Kaplan Meier method. Statistical analysis was performed in IBM SPSS Statistics for Windows, Version 24.0 (25).

### Nucleic acid extraction

DNA and RNA were extracted from histologically confirmed Formalin fixed, paraffin embedded (FFPE) scrolls using the Covaris E220 evolution focused-ultrasonicator and the truTRAC® FFPE total NA Kit. Nucleic acid concentration was quantified using the Qubit™ 3.0 Fluorometer and Qubit™ RNA / DNA HS (high sensitivity) assay kit. Nucleic acid quality was measured by electrophoresis using the Agilent 2200 TapeStation Nucleic Acid System, Agilent 2200 TapeStation Software A.01.05 and the Aligent High Sensitivity RNA / DNA ScreenTape and reagents.

### RNA library preparation, sequencing and bioinformatics

RNA library preparation was performed using the Lexogen Quant Seq 3’ mRNA-Seq Library Prep kit. RNA libraries were denatured, diluted, loaded onto a 75-cycle High output flow cell and sequenced using the NextSeq500 at 2.5-5 million reads (26).

Quality control, trimming and alignment to the reference genome, (NCBI build 37, hg19) was performed with the Partek® Flow® genomics suite software package (Partek, St Louis, MI, USA). The gene expression profiles of primary and CPM and good and poor prognosis cohorts were using gene Specific Analysis (GSA) Modelling using the Partek® flow® with a false discovery rate (FDR) of <0.1.

CMS and CRIS classifications were performed using ‘CMScaller’ (v0.99.1) in the R package, version 2.10.2. (27) (28) Fishers exact test was used to compare contingency between primary and CPM and good and poor prognosis CPM test in IBM SPSS Statistics for Windows, Version 24.0. (25) A p value of < 0.05 was considered significant.

### Methylation array and bioinformatics

DNA was treated with sodium bisulphite using the Zymo EZ-DNA methylation kit, according to manufacturer’s instructions. Degraded FFPE DNA was restored prior to methylation array with the Infinium HD FFPE restore kit, according to manufacturer’s instructions. Methylation array was performed according to the Infinium MethylationEPIC BeadChip Kit manufacturer’s instructions. BeadChips were imaged using the Illumina iScan system. Initial data quality was checked using GenomeStudio Methylation Module Software.

Raw data was loaded into the RStudio version 3.5.0 software using the minifi package. Bioinformatics analysis was performed using the Chip Analysis Methylation Pipeline (ChAMP) R package, version 2.10.2.(29, 30) Probes with signals from less than three functional beads, low confidence with a detection p-value >0.01, covering SNPs, non-CpG and those located on the X and Y chromosome where filtered. Beta-mixture quantile normalization (BMIQ) was applied and a singular value decomposition (SVD) performed to identify batch effects. The association between methylation and prognosis was determined using the Bioconductor R package limma and bumphunter functions. Copy number alteration calling was performed using the CHAMP CNA function with a significance threshold of, p-value < p < ×10^−10^.

### Exome capture, high-throughput sequencing and bioinformatics

DNA was sheared using the Covaris E220 evolution focused-ultrasonicator to produce a uniform 150bp fragment size. Libraries were prepared using the TruSeq® Exome Kit then denatured, diluted, loaded onto a 150-cycle High output flow cell and sequenced using the NextSeq500.

Sequencing reads were assessed using FastQC. Sequences with a Phred score of <30 were removed giving a base call accuracy of 99.9%. Sequence reads were aligned to the human reference genome, (hg19) using the Burrows-Wheeler Aligner (BWA) package (31). SAMTools was used to generate chromosomal coordinate-sorted BAM files and Picard was used to remove PCR duplicates (32). Somatic variants were called from matched tumour-normal samples using Strelka2 in tumour/normal mode (33). Somatic variants were viewed, filtered and annotated in genomics workbench (34). Mutations with a MAF of >1% in known variant databases, (dbSNP and 100,000 genomes) were filtered. Mutations were annotated with information from known variant databases, (dbSNP and 100,000 genomes), PhastCons score and functional consequences. The prognostic groups were compared using Fischer exact test to identify potential candidate driver mutations for poor prognosis CPM. Somatic mutations were entered into the IntOGen platform for further analysis (35). The IntOGen-mutation platform incorporates a number of pipelines to identify cancer driver mutations and activated pathways (35). The OncodriveFM pipeline identifies mutations with a high functional impact using three scoring methods (Sorting Intolerant From Tolerant, (SIFT) (36), PolyPhen2 (37), and Mutation Assessor scores) (35, 38). and assess the likelihood that such mutations are cancer drivers. The OncodriveCLUST pipeline assesses the clustering of mutations to identify relevant activated pathways (35). MSI assessment was carried out using MSI_classifier_v3 (https://rpubs.com/sigven/msi_classification_v3).

## RESULTS

### Patient cohort

From 2011-2017 a total of n=161 patients underwent CRS & HIPEC at University Hospitals Birmingham, n=88 patients for metachronous CPM.

Patients were excluded for the following reasons: rectal primary n=5, incomplete CC2 resection n=8, peritoneal carcinomatosis index (PCI) of ≥ 12 n=20, follow up period of ≤ 12 months n=27 leaving n=28 patients. Complete information regarding the primary CRC pathology and treatment was available for n=26 patients who form the basis of this study. Each patient had matched normal, primary CRC and CPM samples.

Thirteen patients had a DFS of 24 months (15–72 range) following CRS & HIPEC and formed the ‘good prognosis’ cohort, thirteen patients had a DFS of 6 months (2-11 range) and formed the poor prognosis cohort. There were no significant differences between cohorts in demographics, primary CRC or CPM tumour, treatment or follow up (table 1).

**Table 1:** Comparison of good and poor prognosis patient cohorts.

|  |  | **Good**<br>**Prognosis** | **Poor prognosis** | **P value** |
| --- | --- | --- | --- | --- |
| **Age, mean +/- SD** |  | 58 +/-13 | 58 +/-9 | 0.97 |
| **Gender, male** |  | n=7 (54) | n=7 (54) | 0.68 |
| **Tumour**<br>**location** | Right | n=9 (69) | n=6 (46) | 0.33 |
|  | Transverse | n=1 (8) | n=0 (0) |  |
|  | Left | n=3 (23) | n=7 (54) |  |
| **T stage**<br>**primary** | 3 | n=3 (23) | n=3 (23) | 0.66 |
|  | 4a | n=5 (38.5) | n=7 (54) |  |
|  | 4b | n=5 (38.5) | n=3 (23) |  |
| **N stage**<br>**primary** | 0 | n=4 (31) | n=1 (8) | 0.86 |
|  | 1 | n=7 (54) | n=5 (38) |  |
|  | 2 | n=2 (15) | n=7 (54) |  |
| **DFI months** |  | 25 +/-9 | 24 +/-12 | 0.83 |
| **PCI score, median (range)** |  | 5 (3-12) | 8 (2-12) | 0.019 |
| **CC score** | CC0 | n=13 (100) | n=13 (100) | 1 |
|  | CC1 | n=0 (0) | n=0 (0) |  |
|  | CC2 | n=0 (0) | n=0 (0) |  |
| **Follow up, months,**<br>**median (range)** |  | 29 (19-72) | 16 (5-55) | 0.11 |
| **Adjuvant**<br>**treatment** | Yes | n=11 (85) | n=12 (92) | 0.38 |
|  | No | n=2 (15) | n=1 (8) |  |
| **DFS, median (range)** |  | 24 (15-72) | 6 (2-11) | <0.0001 |
| **OS, median (range)** |  | 29 (19-72) | 16 (5-55) | 0.12 |

(N= number value in parenthesis, percentage, DFI, disease free interval, PCI, peritoneal carcinomatosis index, CC score, completeness of cytoreduction, DFS, disease free survival, OS, overall survival)
Log rank $p < 0.0001$ .

Following nucleic acid extraction all patients had adequate CPM RNA for RNAseq (n=13 good, n=13 poor prognosis), n=25 had matched primary CRC samples. For methylation array n=24 patients (n=12 good, n=12 poor prognosis) had adequate DNA. As the Infinium methylation array comprises a 32-prep kit, n=4 good and n=4 poor prognosis primary tumours were matched to these. For exome sequencing n=24 patients (n=12 good, n=12 poor prognosis) had adequate DNA from both the primary and CPM samples, extraction of DNA from normal tissue resulted in n=21 samples (n=9 good, n=12 poor prognosis).

### Exome sequencing

#### Primary tumours

Across all six sequencing runs, we obtained a median of 59.6X coverage (42.3-166.4) with a median uniformity of 88.1% (70.67-89.32). In the primary CRC cohort, after normal subtraction, there was a total of n=112,420 somatic SNV’s, (primary-normal subtraction). Variants present at >1% in the dbSNP common or 1000-Genomes databases were filtered leaving n=38,366 variants.. Seven (7/24, 30%) samples had a high tumour mutational burden, TMB ≥ 10 mut/Mb (39). Mutations were present in 64 of 95 known CRC driver genes (supplementary table S1) (40).

Somatic mutations were entered into the IntOGen pipeline to identify candidate driver mutations. OncodriveFM identified n=46 candidate driver genes with high levels of functional mutation (Q-value<0.05) in the primary CRC cohort (supplementary table S2). The top 10 potential driver genes were *KRAS* (68% samples), *TP53 (*62% samples), *CAD* (40% samples), *APC* (35% samples), *NME1-NME2* (30% samples), *NMD3* and *SDHA* (31% samples), *CDC42BPA* (23% samples) and *ARHGEF12* (23% samples).

#### CPM

In the matched CPM cohort, a total of n=244,531 somatic SNV’s were identified (CPM-primary subtraction). Nine samples, (56%) had a high tumour mutational burden TMB ≥ 10 mut/Mb (39). Mutations were identified in n=69 of n=95 known CRC driver genes, n=51 were shared between the primary and CPM, n=13 were novel (supplementary table S1) (40). Of the somatic variants identified in CPM, n=58,958 (29%) were present in the primary CRC, n=205,552 variants occurred exclusively in the CPM suggesting a significant accumulation of mutations in the transition to CPM (figure 1). OncodriveFM identified n=265 potential driver genes with high levels of functional mutation (Q-value<0.05) in the CPM cohort: *FLNB, SPTB, PPL, TP53, PDE4DIP, RIOK2, CDC16, NUP98, CDC16* and *SVEP1* (supplementary table S2), however these results must be treated with caution due to the bias of the hypermutator phenotype. KEGG pathway analysis of mutations demonstrated enrichment in pathways concerning the immune system, signalling, metabolism and cancer (supplementary table S3).

**Figure 1:**
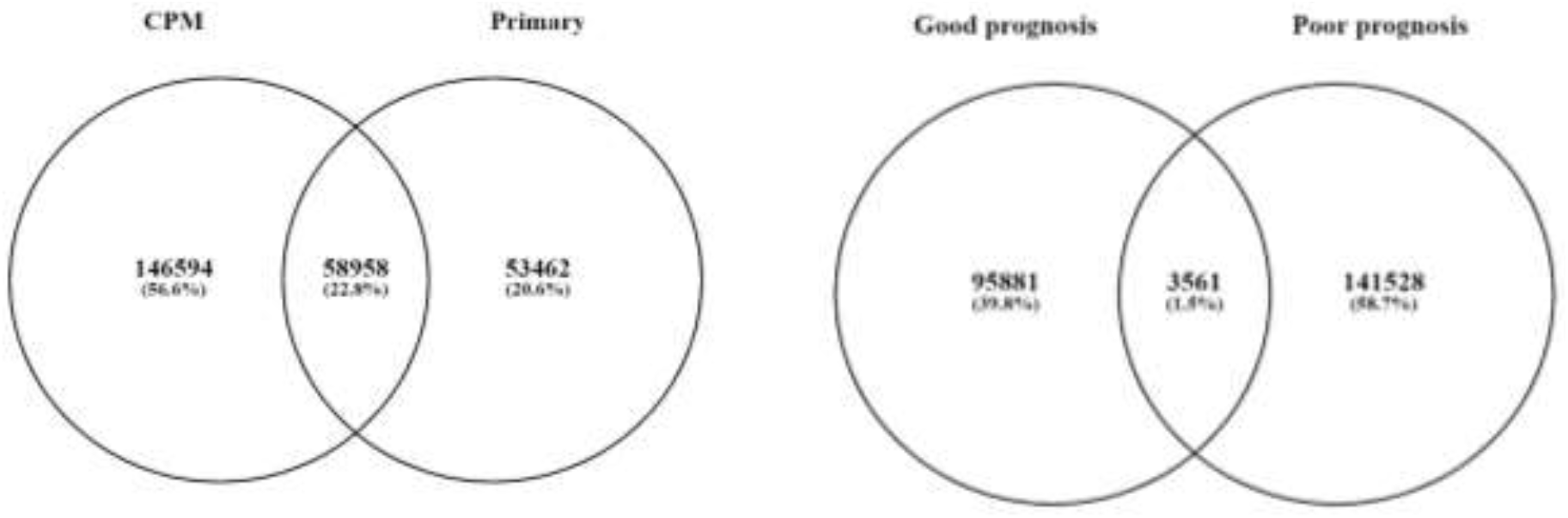
Mutations exclusive to and shared between primary CRC and matched CPM and good and poor prognosis CPM

Clonality analysis with SuperFreq showed distinct differences between the good and poor prognosis groups, with a median of 2 clones in the good prognosis group of primary tumours (range 1-4) and 3 clones in the poor prognosis group (range 2-7). In the peritoneal metastases there were a median of 3 clones in both the good (range 1-4) and poor (range 2-5) groups. Of note, in the poor prognosis group during clonal expansion, the dominant clone in the peritoneal metastasis group arose de-novo rather than being a prior clone that existed in the primary tumour (Supplementary figure 1).

In the primary tumours, 4/21 (19.0%) were MSS and 17/21 MSI (81.0%) whereas in the isolated peritoneal metastasis group, 9/21 were MSI (47.4%) and 10/21 were MSS (52.6%) demonstrating that there was a significantly higher rate of MSI in the isolated peritoneal metastasis group (p<0.05, Chi2).

Patients with poor prognosis CPM had a higher frequency of somatic mutations: 60% of all mutations in CPM cohort vs. 40%. Patients with poor prognosis more commonly had a high tumour mutational burden, TMB ≥ 10 mut/Mb (39), 56% vs. 44%. Of the somatic mutations identified in poor prognosis CPM, n=35,461 (30%) were present in patients with good prognosis CPM, n=145,089 variants occurred exclusively in patients with poor prognosis CPM, suggesting a high tumour mutational burden was associated with the development of poor prognosis CPM (figure 1).

Comparison of somatic mutations in patients with good and poor prognosis identified two potential candidate genes, *FAM13A* and *PIEZO2* (Fishers exact p<0.05, FDR=0.53) (table 2).

**Table 2.** Potential candidate variants, poor prognosis CPM.

| Chr | Position | Reference | Allele | P-value | FDR | Sample frequency (case) | Sample frequency (control) | Gene ID |
| --- | --- | --- | --- | --- | --- | --- | --- | --- |
| 4 | 93084410 | C | G | 0.007 | 0.53 | 62.5 | 0 | FAM13A |
| 18 | 11552313 | G | C | 0.023 | 0.53 | 50 | 0 | PIEZO2 |

CPM identified through Fisher exact test, genomics workbench (Chr, chromosome, FDR, false discovery rate)

### Gene expression

#### Differences between primary CRC and matched CPM

Primary CRC and matched CPM showed differential expression of n=65 genes with an FDR <0.1. (Figure 2) Sixteen genes showed significantly decreased expression in CPM compared with primary CRC (table 3). Forty-nine genes showed significantly increased expression in CPM compared with primary CRC (table 3). A KEGG pathway analysis was performed to identify the enriched biological functions among the differentially expressed genes (table 4). The expression of *FABP6*, an intercellular bile acid transporter, was decreased 34.30-fold in CPM. *OLFM4* is a target of the Wnt/β-catenin pathway, its expression was reduced 3.77-fold in CPM. *DCN* and PTEN ar able to initiate a number of signalling pathways including *ERK* and *EGFR* leading to growth suppression, their expression was increased 3.3-fold and 3.25 fold in CPM, this was unexpected and in contrast to the literature. (41). *NF-κBIA* expression was increased 3.24-fold in CPM, its upregulation may reflect increased *NF-κB* activity in the development of CPM (42).

**Table 3.** The top 10 genes with significantly altered expression (FDR<0.1) in CPM samples compared with primary CRC samples.

| Rank | Gene name | Function | Fold change | FDR p value |
| --- | --- | --- | --- | --- |
| **Reduced expression CPM samples vs. primary CRC** |  |  |  |  |
| 1 | FABP6 | Intracellular bile acid transporter | -34.30 | 1.74 x 10<sup>-06</sup> |
| 2 | DEFA6 | Cytotoxic peptide involved in host intestine defence | -8.15 | 8.55 x 10<sup>-06</sup> |
| 3 | DMBT1 | Tumour suppressor | -6.06 | 2.43 x 10<sup>-04</sup> |
| 4 | TTC38 | Protein coding gene | -4.56 | 5.80 x 10<sup>-05</sup> |
| 5 | OLFM4 | Wnt/β-catenin pathway target | -3.77 | 1.01 x 10<sup>-04</sup> |
| 6 | IGHA1 | Immune receptor | -3.66 | 4.23 x 10<sup>-05</sup> |
| 7 | CES2 | Intestinal enzyme controlling drug clearance | -3.20 | 6.84 x 10<sup>-05</sup> |
| 8 | NDUFS6 | Enzyme in electron transport chain of mitochondria | -2.70 | 7.74 x 10<sup>-05</sup> |
| 9 | P2RY11 | G-protein coupled receptor | -2.53 | 6.37 x 10<sup>-04</sup> |
| 10 | MUC2 | Encodes a mucinous intestinal coating | -2.34 | 7.22 x 10<sup>-04</sup> |

| Increased expression CPM samples vs. primary CRC |  |  |  |  |
| --- | --- | --- | --- | --- |
| 1 | CD53 | Tetraspanin | 7.29 | 5.87 x 10<sup>-05</sup> |
| 2 | CYR61 | Extracellular signalling protein | 4.24 | 3.12 x 10<sup>-04</sup> |
| 3 | CXCL12 | G-protein coupled receptor | 3.64 | 9.25 x 10<sup>-04</sup> |
| 4 | NR2F1 | Nuclear hormone receptor and transcriptional regulator | 3.53 | 7.09 x 10<sup>-04</sup> |
| 5 | CTGF | Connective tissue growth factor | 3.49 | 1.55 x 10<sup>-04</sup> |
| 6 | CSTB | Cystatin | 3.41 | 6.13 x 10<sup>-04</sup> |
| 7 | TSC22D3 | Anti-inflammatory protein glucocorticoid (GC)-induced leucine zipper | 3.36 | 3.94 x 10<sup>-04</sup> |
| 8 | DCN | Tumour suppressor gene | 3.30 | 6.19 x 10<sup>-05</sup> |
| 9 | PTEN | Tumour suppressor gene | 3.25 | 9.28 x 10<sup>-04</sup> |
| 10 | NF-κBIA | Inhibits the *NF-κB* transcription factor | 3.24 | 1.06 x 10<sup>-04</sup> |

**Table 4.** CMS and CRIS classification good vs. poor prognosis CPM.

|  | **Non-consensus** | **CMS1** | **CMS2** | **CMS3** | **CMS4** |  | **Total** |
| --- | --- | --- | --- | --- | --- | --- | --- |
| **Good prognosis CPM** | 10 (77) | 0 (0) | 0 (0) | 1 (8) | 2 (15) |  | 13 |
| **Poor prognosis CPM** | 2 (15) | 1 (8) | 3 (23) | 1 (8) | 6 (46) |  | 13 |
|  | **Non-consensus** | **CRISA** | **CRISB** | **CRISC** | **CRISD** | **CRISE** |  |
| **Good prognosis CPM** | 5 (39) | 2 (15) | 2 (15) | 3 (23) | 0 (0) | 1 (8) | 13 |
| **Poor**<br>**prognosis**<br>**CPM** | 1 (8) | 2 (15) | 5 (39) | 2 (15) | 3 (23) | 0 (0) | 13 |

*CMS Fishers exact p-value 0.005, CRIS Fischer's exact p-value 0.148, values in parenthesis percentages*

**Figure 2:**
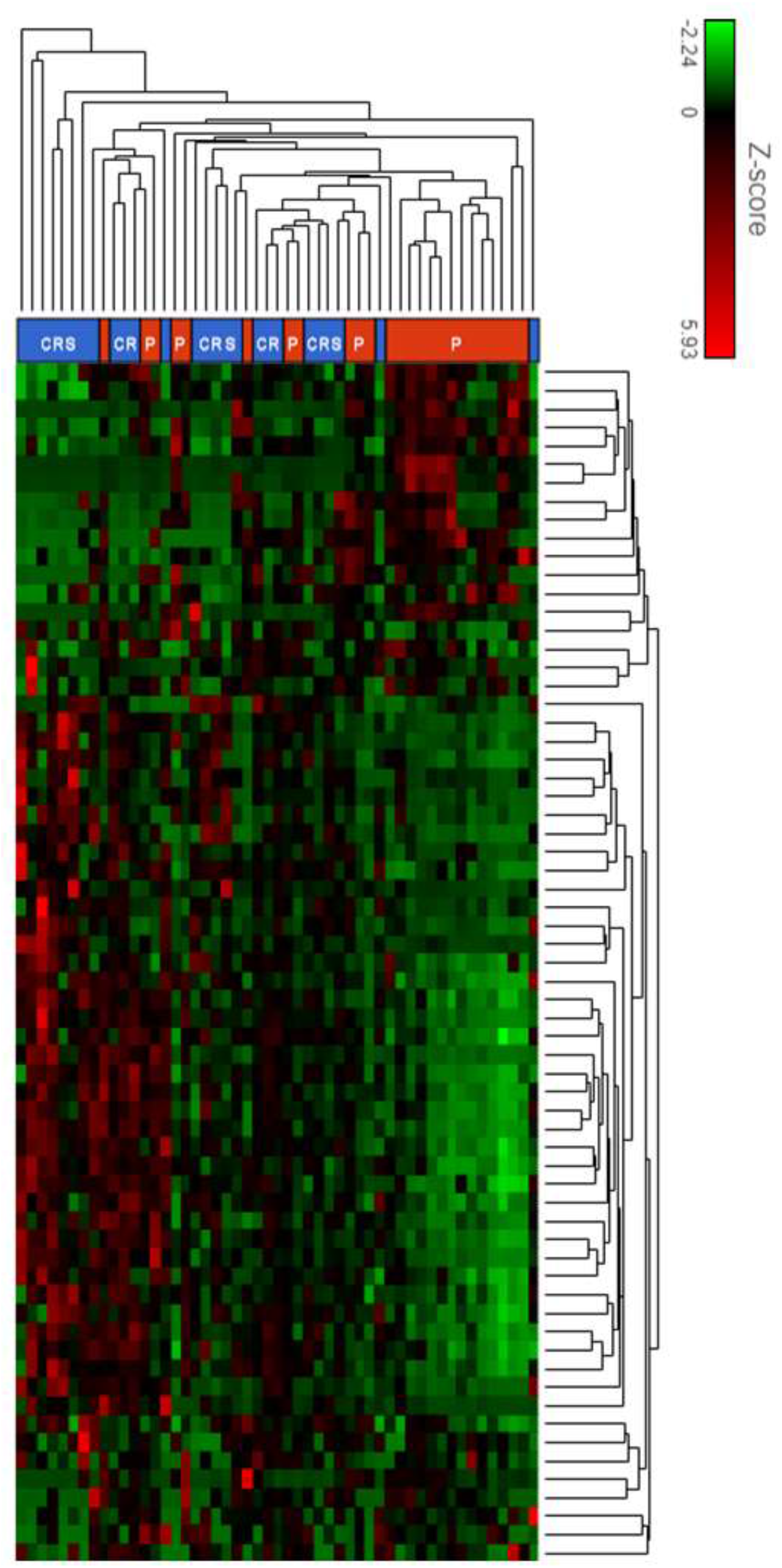
Heatmap of differential gene expression between CPM vs. primary samples. Sample type is indicated at the upper border of the heatmap. Blue CRS = CPM; P red=primary

#### Differential gene expression profile Poor vs. Good prognosis CPM

One hundred and fifty-nine genes showed increased expression in poor prognosis CPM (table 5, figure 3). Five genes showed decreased expression in in poor prognosis CPM, however none had a fold change ≥ 1.5 suggesting minimal difference in expression between the good and poor prognosis cohorts (table 5). KEGG pathway analysis demonstrated enrichment in endocytosis, metabolism, phagocytosis, cell movement and architecture, bacterial and viral cell infection, transcription and the expression of genes controlling apoptosis, cell cycle, oxidative stress resistance and longevity (table 3). The expression of *CEACAM1*, a member of the carcinoembryonic antigen (*CEA*) immunoglobulin family, was increased 8.27-fold in poor prognosis CPM (43).

**Figure 3:**
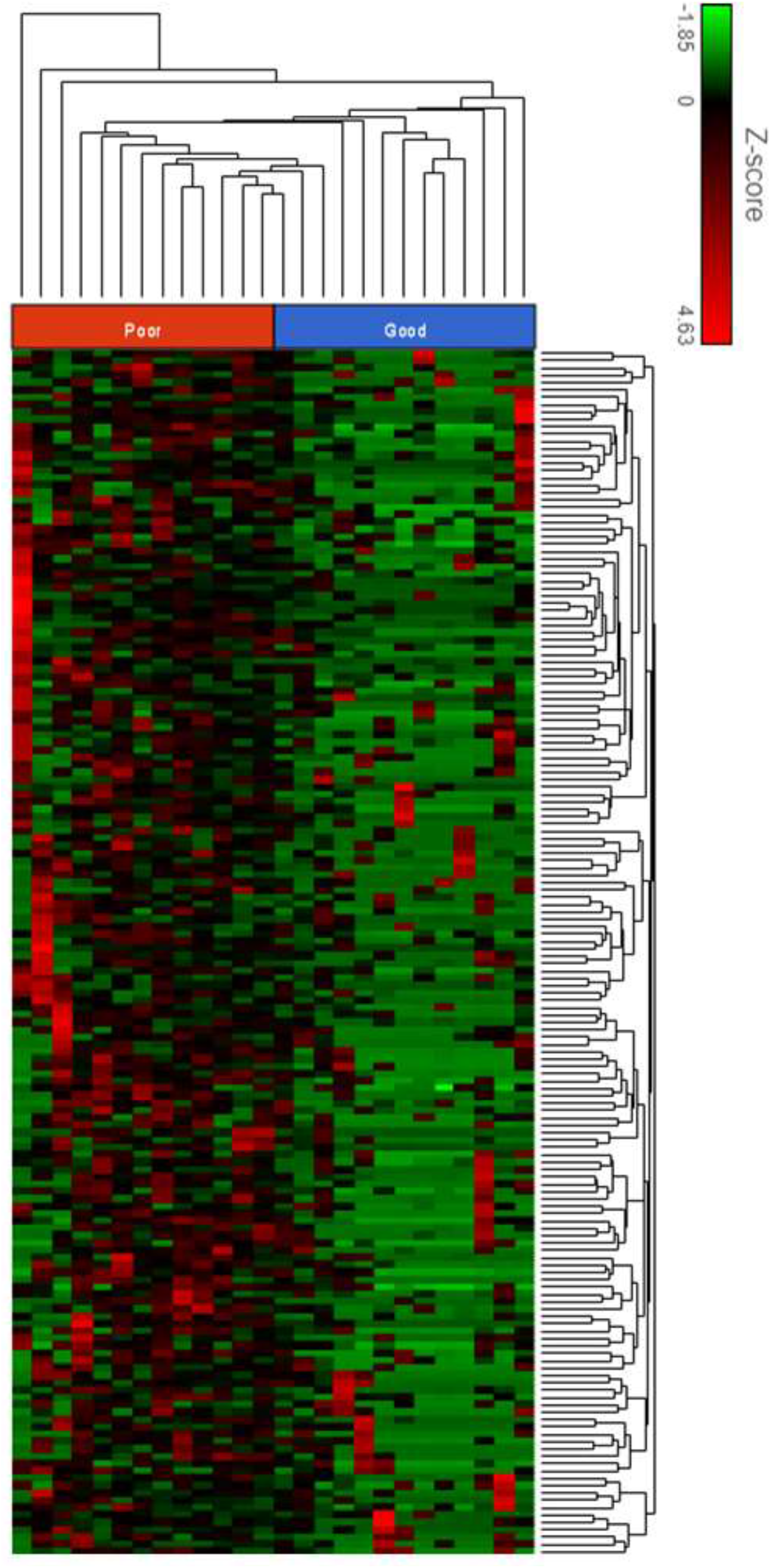
Heatmap differential gene expression good vs. poor CPM. Sample type is indicated at top of heatmap. Poor = poor prognosis; Good = good prognosis.

*AXIN1* encodes a cytoplasmic protein which forms the ß-Catenin destruction complex, a negative regulator of the WNT signalling pathway (44). *AXIN1* expression was increased 5.42-fold in poor prognosis CPM which suggests increased activity of the Wnt signalling pathway (45) and may suggest sensitivity to porcupine inhibition.

Amongst the n=51 primary CRC and CPM samples n=35 were representative of each CMS subtype, the remaining n=16 samples did not have a consistent pattern (figure 4). Comparison of the CMS subtypes in primary and CPM and prognostic groups revealed an apparent transition from primary CRC to CPM. No primary CRC samples were classified as CMS4 (mesenchymal subtype characterized by prominent transforming growth factor activation, stromal invasion and angiogenesis) compared to 31% of CPM (p=0.085). Secondly, patients with poor prognosis CPM were more commonly CMS4, 46% vs. 15% (p=0.005, table 6).Amongst the n=51 primary CRC and CPM samples n=42 were representative of each CRIS subtype, the remaining nine samples did not have a consistent pattern (figure 5). There was no significant difference in CRIS classification between primary and CPM, p-value 0.365, or between good and poor prognosis CPM, p-value 0.148, (table 6). The non-concordance with CMS subtypes may reflect the increased stromal mesenchymal tissue present in patients with CPM and particularly those with CPM. As CRIS classification is based only on tumour cells such changes in intratumoural heterogeneity are not accounted for.

**Figure 4:**
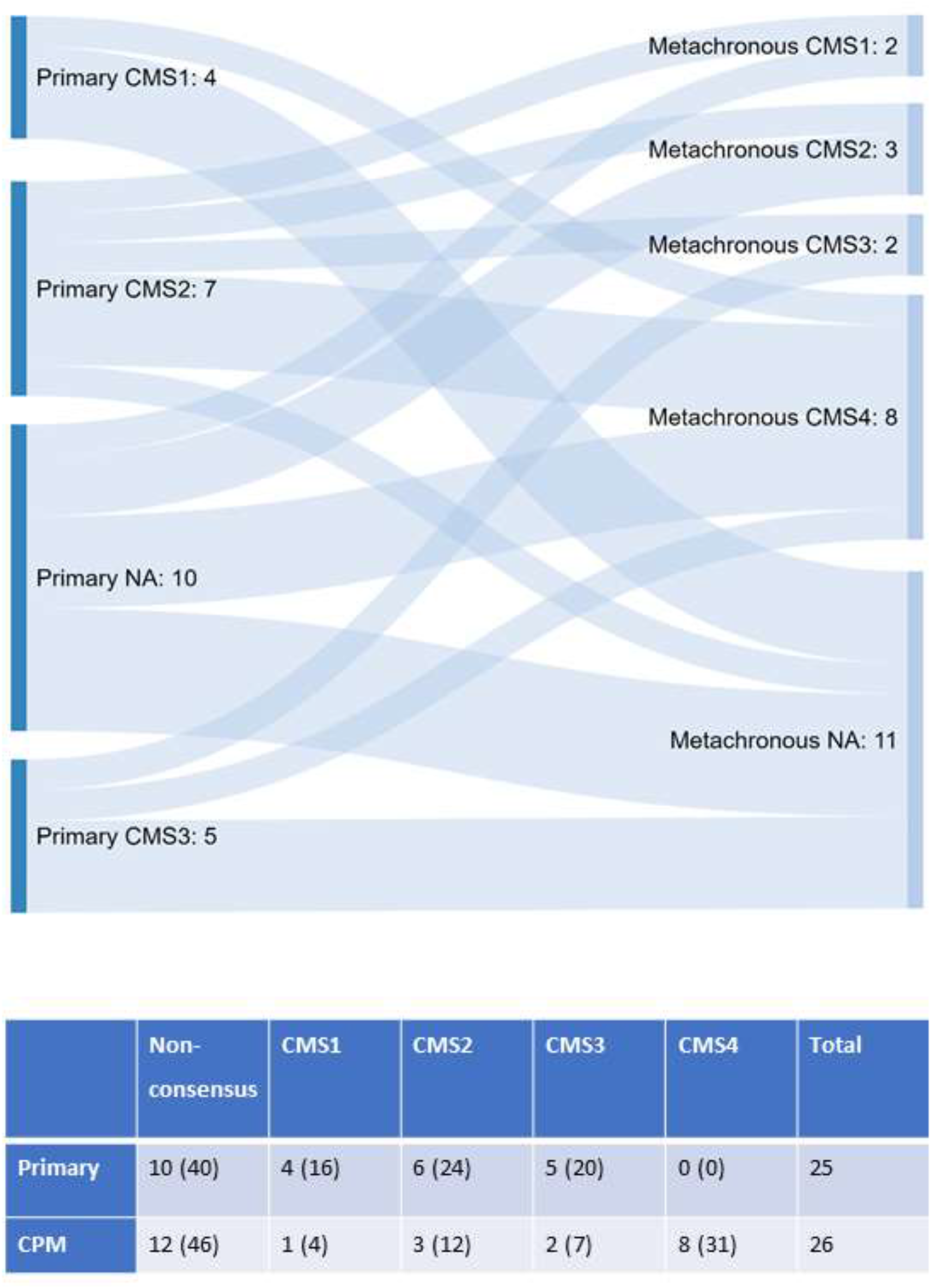
Sankey diagram of the CMS transition from primary to CPM and CMS classification. Fischers exact p-value 0.085, values in parenthesis percentages

**Figure 5:**
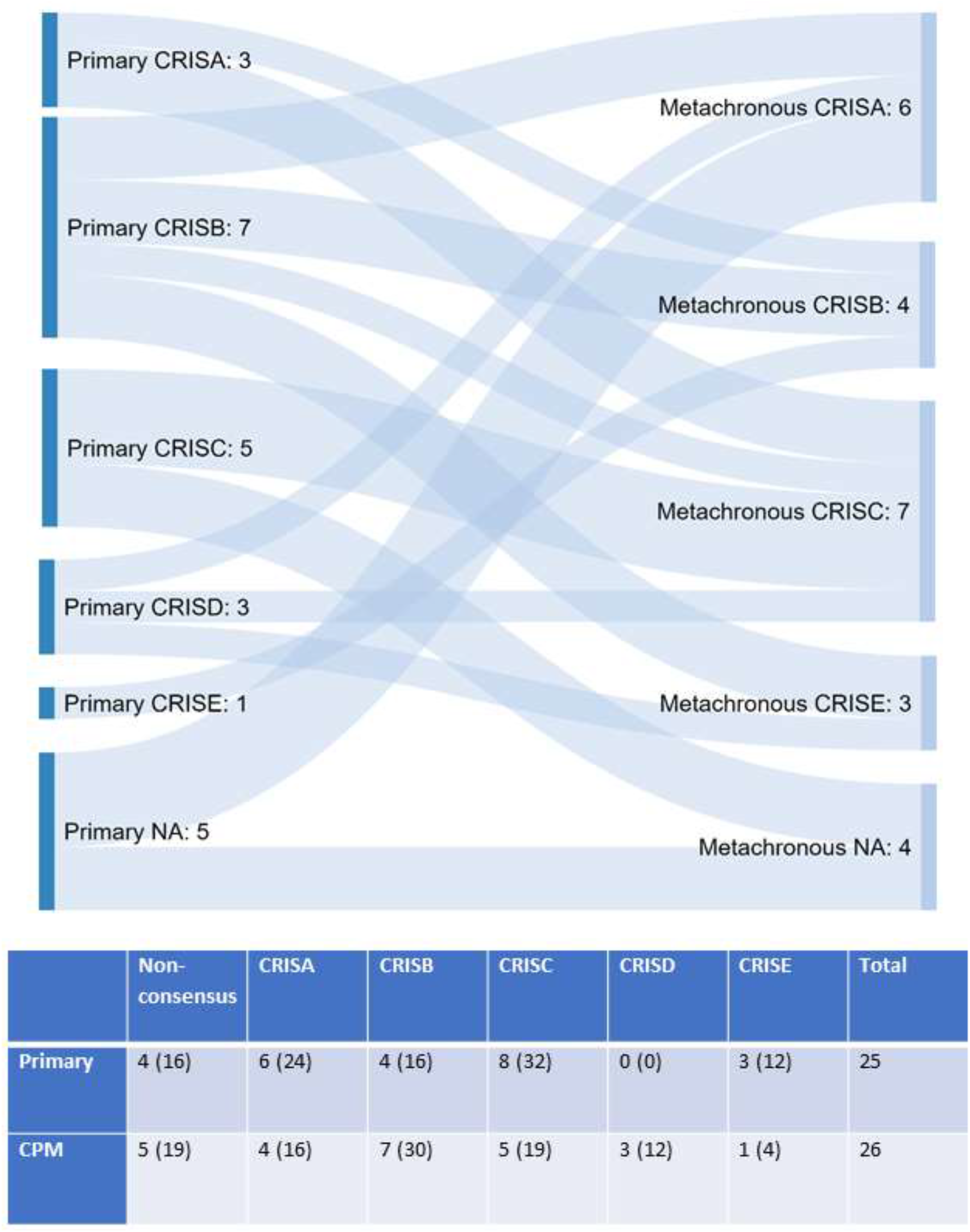
Sankey diagram of the CRIS transition from primary to CPM. Fishers exact p-value 0.365, values in parenthesis percentages

### Methylation

#### Methylation Microarray Analysis, primary CRC vs. matched metachronous CPM

Thirty-two samples in total were hybridised successfully to the Illumina HumanMethylation EPIC microarrays. DMPs were called between the primary CRC and CPM. The top ranked differentially methylated probe was cg04146982, BF 34.5, adjusted p-value 5.67 × 10-16 (chr8:144,943,810-144,943,810, hg19 coordinates), which tags a CpG dinucleotide 3651 bp upstream of the transcription start site of gene Epiplakin 1, (EPPK1) (46). EPPK1 is part of the Plakin family an important component of the cell cytoskeleton (47). The other DMP was cg12209861, BF 7.1, adjusted p-value 0.059 (chr4:37,459,078-37,459,078, hg19 coordinates), 3526 bp upstream of the transcription start site of gene Chromosome 4 Open Reading Frame 19, (*C4orf19*). DMRs were called between primary CRC and CPM via the dmrLasso function of the CHAMP pipeline (table 7). The top 10 most significant DMRs were in the region of *IGF2, ZNF461, RASGFR1, CELF4, ZSCAN18, EDNRB, ZBED9, VTRNA2-1, ZNF256 and EGFLAM*. KEGG pathway analysis did not reveal any significantly enriched pathways.

Comparison of CNA between primary and CPM via methylation arrays did not identify and significant differences in CNA between primary and CPM at a stringent p-value of < ×10^−10^ however a number of CNA were identified at a lower significance threshold, p=2.78 ×10−^07^ (table 8).Genes showing CNA gains of known significance in patients with CPM included; *TRIM3, 5, 6, 21 and 22, MT1A, 2A, 3, 4* encode proteins of the metallothionein family.

#### Methylation Microarray Analysis, Poor vs. Good prognosis CPM

The top ranked differentially methylated probe was cg07951355, BF=6, (chr1:40123717) which tags an intergenic region 1076 bp before gene *NT5C1A*. Cg25909064, BF 4 adjusted p-value 0.47 (chr11:120081487-120082345) which tags an intron of gene *OAF* and cg12977942, BF 4 adjusted p-value 0.47 (chr5:92839309-92839309) which tags an intron of gene *NR2F1-AS1* (46). Six significant DMRs (table 7) were identified in the regions of *NKX6-2, CHFR, GATA3. IRX5* and *HCK* and *BC019904*. KEGG pathway analysis did not reveal any significantly enriched pathways.

Comparison of CNA between the CPM prognostic groups identified recurrent gene losses at chromosomes 3, 4, 14, 15, 17 and 19 (table 8). CNA losses clustered in the RAS-MAPK-ERK signalling pathway suggesting dysregulation in poor prognosis CPM. Comparison of CNA between the CPM prognostic groups identified n=19 gene gains at chromosomes 9, 10 and 11. Genes showing CNA gains in patients with poor prognosis CPM included: *SIT1, RNF38, MELK, PAX5, SHB, ZEB1, DEAF1, ANTXR, EPS8L2* and *PIDD1*

## DISCUSSION

This study determined the gene expression, CNA, methylation and somatic mutation profile of primary CRC and matched isolated CPM to determine whether there were a change associated with the development of CPM or predicting prognosis for patients with CPM. To our knowledge, this is the first such analysis in a cohort of patients with isolated CPM suitable for treatment with CRS & HIPEC. The MSKCC cohort of metastatic cancer (23) had a diverse range of metastatic cancer, none of whom overlapped with the type we have studied, which is isolated colorectal peritoneal metastasis, with matched primary samples, suitable for cytoreduction.

Comparison of patients with primary CRC and metachronous CPM identified biological changes associated with the transition from primary CRC to CPM. Hypermethylation, CNA and hypermutation resulted in the inactivation of tumour suppressors and oncogene activation in CPM, (TP53, VTRA2-1, TRIM proteins). These changes suggest a rapid rate of tumour growth unchecked by tumour suppressor or apoptotic mechanisms.

Increased MAPK and Wnt/β-catenin pathway activation was noted in CPM. Gene expression of negative regulators of the Wnt pathway was reduced, (OLFM4, DEAFA6), negative Wnt regulators contained somatic mutations, (APC, RNF43, FAM123B and TSC1), and the MAPK marker, RASFGFR1 was hypermethylated suggesting persistent activation of MAPK and Wnt pathways. Multiple mutations of negative Wnt signalling regulators make this an attractive therapeutic target. Porcupine inhibitors mediate the palmitoylation of Wnt ligands, blocking Wnt signalling. The porcupine inhibitor LGK974 inhibits the upstream negative Wnt regulator mutant RNF43 and is a potential therapeutic target in CPM (48).

CPM contained a high proportion of MSH6 somatic mutations suggesting deficiency in the mismatch repair pathway and MSI. MSH6 mutations are commonly found in isolated peritoneal metastasis (49). As expected for tumours with mismatch repair deficiency both the primary CRC and CPM cohort had a high tumour mutational burden, crucially this suggests they may have a good response to treatment with immune checkpoint inhibitors such as pembrolizumab (50), a new therapeutic avenue for these difficult to treat patients. The frequency of hypermutatation seen in our study (48%) was considerably higher than that observed for both the MSKCC metastatic disease cohort (5%) and the TCGA Colorectal (51) cohort (10%). The expression of genes regulating innate immunity however was downregulated, (DEFA6, DMBT1, MUC2) or altered via somatic mutations, (HLA-A antigen) suggesting immune evasion in the transition to CPM which may reduce the likelihood of successful PD-1 therapy.

The expression of genes supressing invasion, migration and EMT was downregulated or hypermethylated, (MUC2, MMP26, ILK, FLNB, SPTB, PPL, and SVEP1) and those triggering these processes upregulated, (CYR61, CXCL12, CTGF, CSTB). These changes suggest a mechanism by which CPM cells metastasise from the primary CRC. In keeping with changes in EMT regulators there appeared to be a transition in CMS subtypes towards CMS4 from primary CRC to CPM. The CMS4 subtype is an interesting therapeutic target, *TGFβ* signalling inhibitors and targeted immunotherapies have been trialled with success in pre-clinical models to block cross talk between the tumour microenvironment and halt disease progression of stromal rich CMS4 CRC (52, 53).

Methylation appeared to be dysregulated in CPM with a bias towards a hypermethylator phenotype caused by somatic mutation of the TET2 tumour suppressor and CDH7 chromatin regulator. Active DNA demethylation by TET enzymes is an important tumour suppressor mechanism in a variety of cancers (54-56). Downregulation of CES2, a gene known to activate the prodrug irinotecan, a commonly used chemotherapy in the adjuvant treatment of primary CRC and CPM was seen in this cohort. Resistance to the treatment of primary CRC may in part explain the development of CPM.

*CEACAM1* expression correlates with metastasis and reduced survival in CRC and was upregulated in this cohort of patients (57). Novel therapies in the form or CEA TCB IgG-based T-cell bispecific antibodies (Cibisatamab) may therefore be of benefit (58). Additionally there was a downregulation of gene expression of negative regulators of the Wnt pathway, (AXIN1) and somatic mutations of key Wnt regulators, (FAM13A) and hypermethylation of MAPK and TGF-β pathway markers, (RAB8A, RAB34, FGF5 and BMP3) suggesting persistent activation of MAPK, TGF-β and Wnt in poor prognosis CPM.

A relative weakness of this study is the small cohort of patients, the biological changes identified here form a starting point in identifying the tumour biology associated with the development of CPM and predicting poor prognosis disease. However, we have identified multiple potential targets for therapy, along with the important finding that CPM appears to be a hypermutated, hypermethylated, immune evasive cancer which allows it to be potentially targeted by emerging novel therapeutics.

Patients with colorectal peritoneal metastasis (CPM) secondary to colorectal cancer have limited survival with the best available treatments. Despite selection for treatment using known prognostic factors survival varies widely and can be difficult to predict. There is a paucity of knowledge concerning the biology of CPM, it is likely that there are additional biological markers of response to currently available as well as novel or re-purposed alternative treatments. Here we have comprehensively profiled a cohort of patients with isolated CPM and identified a number of therapeutically targetable alterations including mutations in Wnt/β catenin regulators, the mismatch repair pathway and methylation regulators.

## Data Availability

Data will be deposited onto SRA on acceptance of manuscript

## DATA AVAILABILITY

The data that support the findings of this study are available from the corresponding author upon reasonable request.

## Supplementary tables & figures

**Supplementary figure 1:**
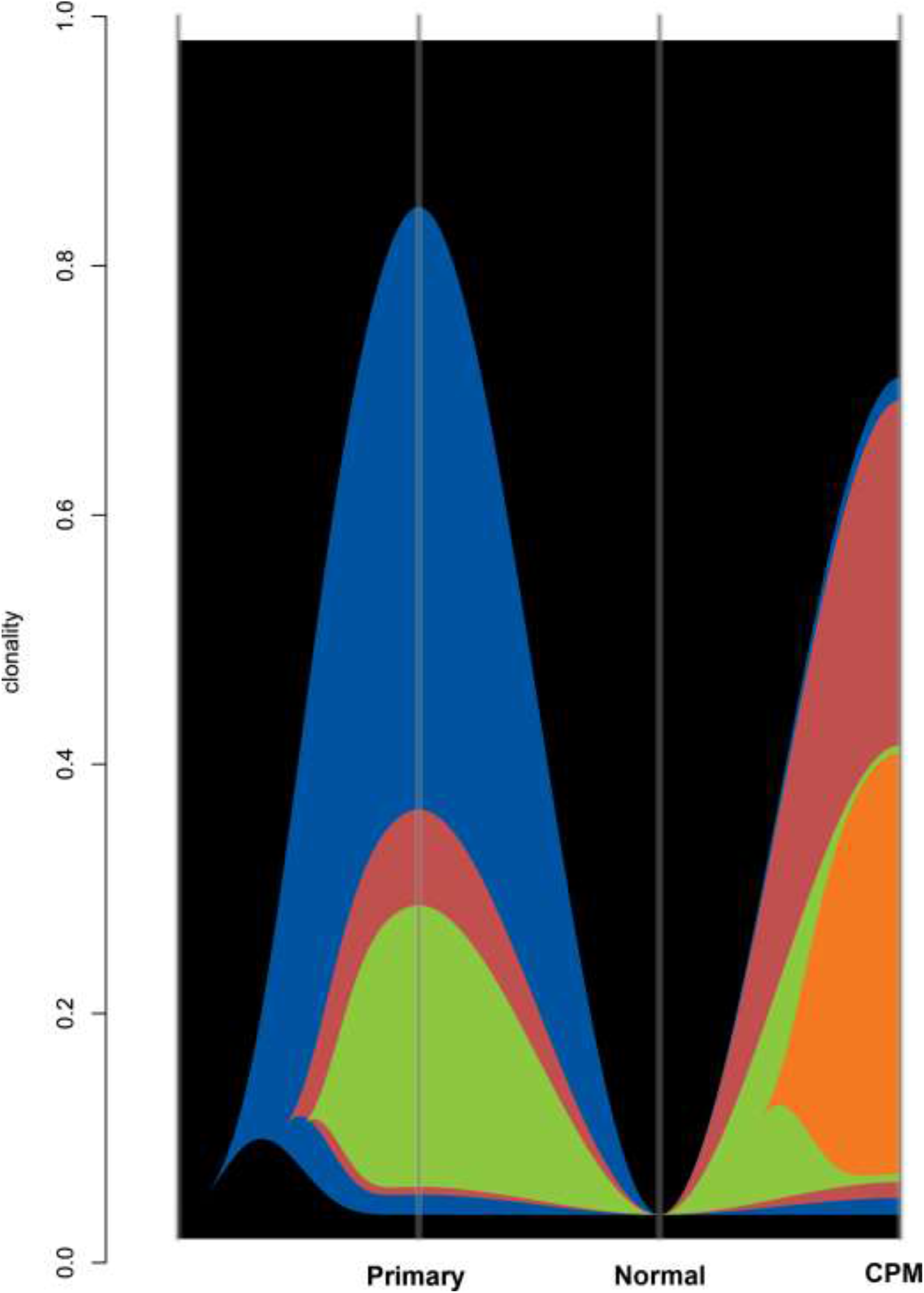
Clonality river plot of primary tumour, normal germline and colorectal peritoneal metastasis (CPM). Clonality on Y axis.

**Supplementary table 1:** KEGG pathway enrichment analysis, primary CRC vs. matched metachronous CPM and good vs. poor prognosis CPM (p-value < 0.05)

| **Gene set** | **Class** | **KEGG pathway** | **Enrichment score** | **P-value** |
| --- | --- | --- | --- | --- |
| **Primary CRC vs. matched CPM** |  |  |  |  |
| hsa04810 | Cellular Processes; Cell motility | Regulation of actin cytoskeleton | 4.19 | 0.02 |
| hsa05206 | Human Diseases; Cancers: Overview | MicroRNAs in cancer | 3.44 | 0.03 |
| hsa05222 | 'Human Diseases; Cancers: Specific types | Small cell lung cancer | 3.43 | 0.03 |
| hsa04672 | Organismal Systems; Immune system | Intestinal immune network for IgA production | 3.30 | 0.04 |
| hsa04670 | Organismal Systems; Immune system | Leukocyte transendothelial migration | 3.22 | 0.04 |

| Good vs. Poor prognosis CPM |  |  |  |  |
| --- | --- | --- | --- | --- |
| hsa04144 | Cellular Processes;<br>Transport and<br>catabolism | Endocytosis | 11.67 | 8.56<br>x10-6 |
| hsa00130 | Metabolism;<br>Metabolism of cofactors<br>and vitamins | Ubiquinone and<br>other terpenoid-<br>quinone<br>biosynthesis | 5.07 | 6.28<br>x10-3 |
| hsa05100 | Human Diseases;<br>Infectious diseases:<br>Bacterial | Bacterial invasion<br>of epithelial cells | 4.76 | 8.56<br>x10-3 |
| hsa05110 | Human Diseases;<br>Infectious diseases:<br>Bacterial | Vibrio cholerae<br>infection | 4.14 | 0.02 |
| hsa04145 | Cellular Processes;<br>Transport and<br>catabolism | Phagosome | 3.92 | 0.02 |
| hsa05165 | Human Diseases;<br>Infectious diseases:<br>Viral | Human<br>papillomavirus<br>infection | 3.85 | 0.02 |
| hsa00561 | Metabolism; Lipid<br>metabolism | Glycerolipid<br>metabolism | 3.64 | 0.03 |
| hsa04966 | Organismal Systems;<br>Excretory system | Collecting duct<br>acid secretion | 3.63 | 0.03 |
| hsa00230 | Metabolism; Nucleotide<br>metabolism | Purine metabolism | 3.37 | 0.03 |
| hsa05120 | Human Diseases;<br>Infectious diseases:<br>Bacterial | Epithelial cell<br>signalling in<br>*Helicobacter pylori*<br>infection | 3.31 | 0.04 |
| hsa00020 | Metabolism;<br>Carbohydrate<br>metabolism | Citrate cycle (TCA<br>cycle) | 3.20 | 0.04 |
| hsa01230 | Biosynthesis of amino<br>acids | Biosynthesis of<br>amino acids | 3.20 | 0.04 |
| hsa04520 | Cellular Processes;<br>Cellular community –<br>eukaryotes | Adherens junction | 3.13 | 0.04 |
| hsa03020 | Genetic Information<br>Processing;<br>Transcription | RNA polymerase | 11.67 | 8.56<br>x10-6 |
| hsa04068 | Environmental<br>Information Processing;<br>Signal transduction | FoxO signalling<br>pathway | 5.07 | 6.28<br>x10-3 |

**Supplementary table 2:**
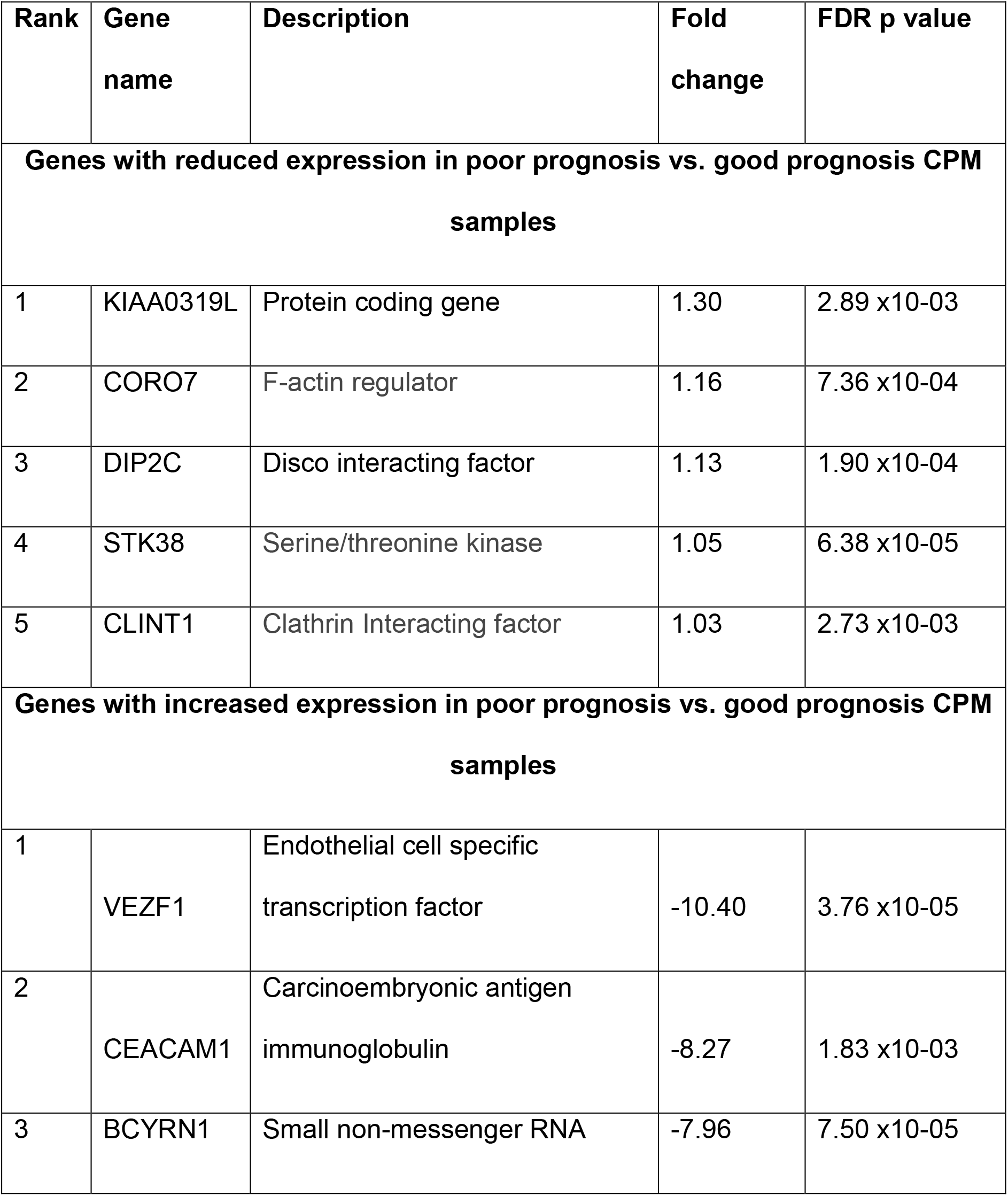
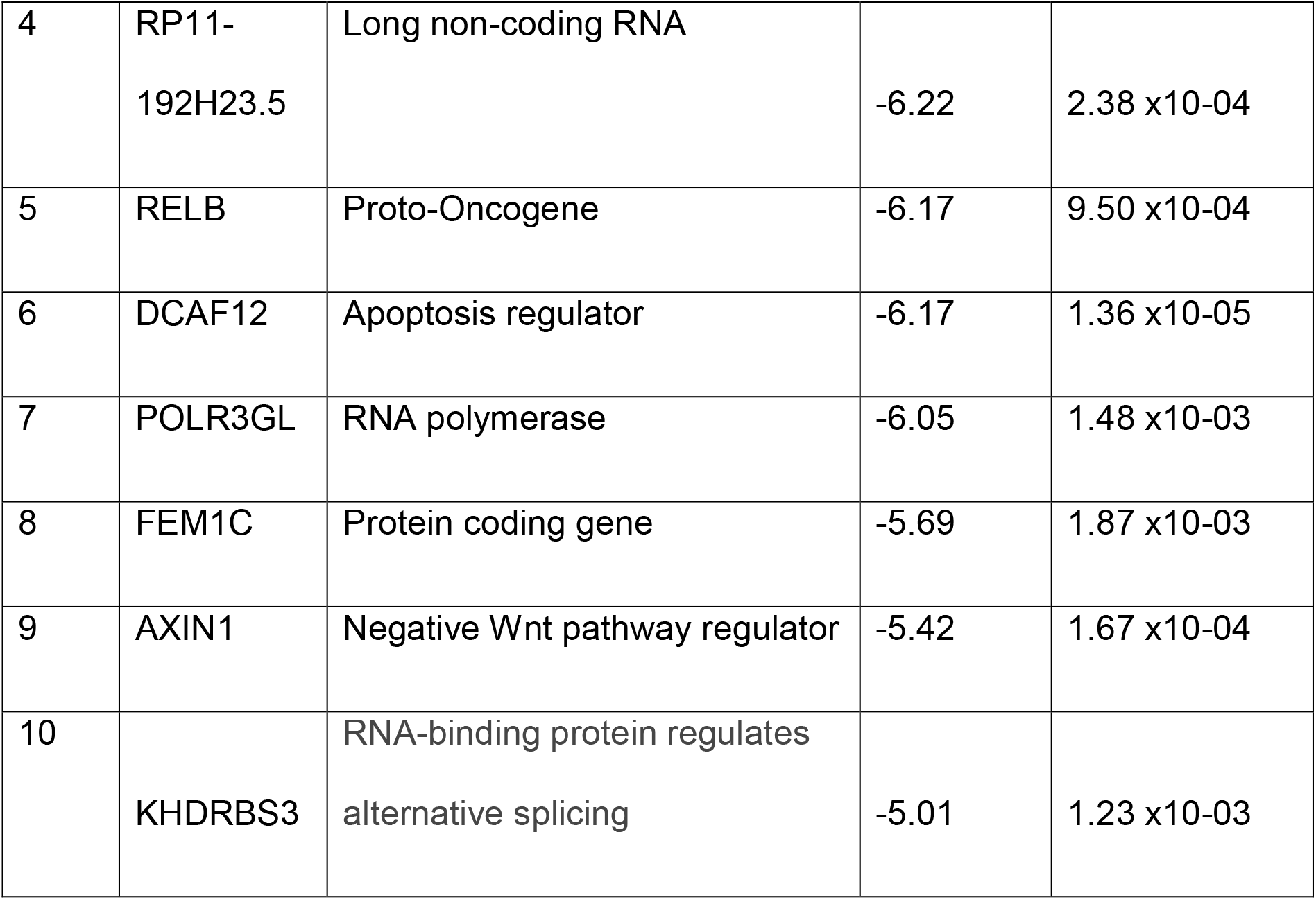
The top 10 genes with significantly altered expression (FDR<0.1) in poor prognosis CPM.

**Supplementary table 3:**
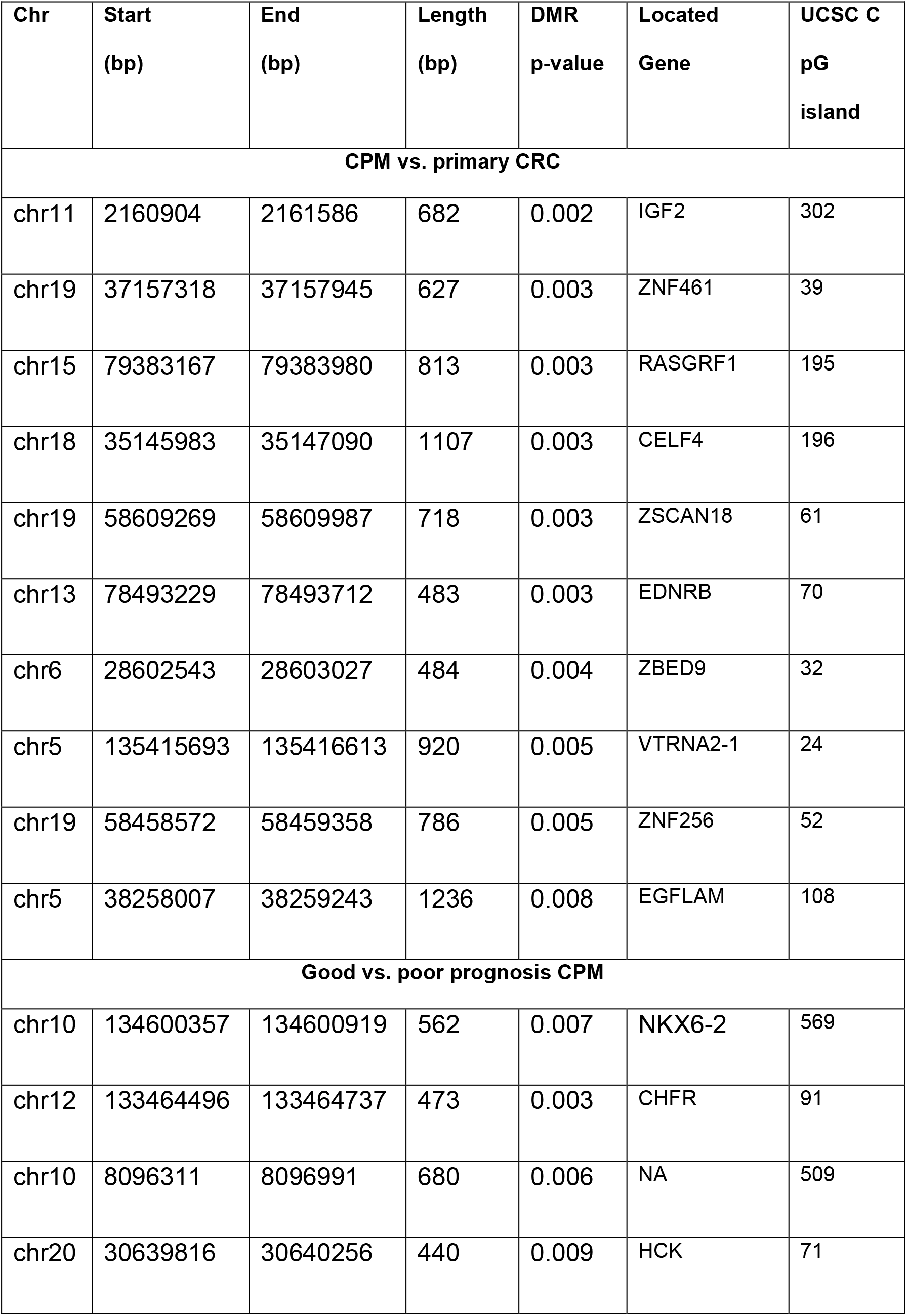
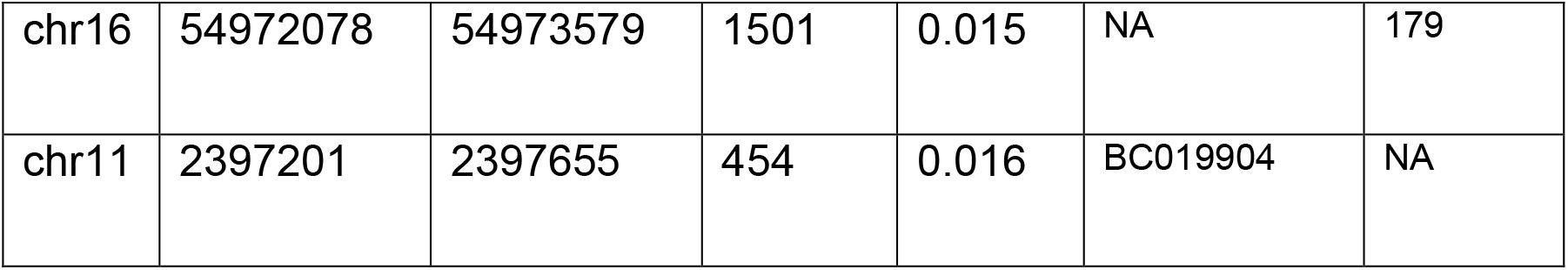
The top 10 differentially methylated regions, (DMRs), CPM vs. primary CRC and good vs. poor prognosis CPM.

**Supplementary table 4:** The top 10 CNAs, CPM vs. primary CRC and good vs. poor prognosis CPM.

| Chr | Start | End | Type | Count | Score | GW_P | Gene |
| --- | --- | --- | --- | --- | --- | --- | --- |
| **CPM vs. primary CRC** |  |  |  |  |  |  |  |
| chr11 | Gain | 3817692 | 3818253 | 22 | 0.91 | 2.78<br>X10-07 | NUP98 |
| chr11 | Gain | 3825449 | 4629195 | 22 | 0.91 | 2.78<br>X10-07 | RRM1,<br>TRIM21 |
| chr11 | Gain | 4629529 | 5704417 | 22 | 0.91 | 2.78<br>X10-07 | MMP26,<br>UBQLN3,<br>TRIM5, 6 |
| chr11 | Gain | 5705995 | 5705995 | 22 | 0.91 | 2.78<br>X10-07 | TRIM5 |
| chr11 | Gain | 5712109 | 6518263 | 22 | 0.91 | 2.78<br>X10-07 | TRIM3, 5,<br>22,<br>FAM160A2,<br>CCKBR,<br>SMPD1,<br>APBB1,<br>ARFIP2 |
| chr11 | Gain | 6518907 | 6650480 | 22 | 0.91 | 2.78<br>X10-07 | DNHD1,<br>ILK, TPP1,<br>TAF10,<br>RRP8 |
| chr16 | Gain | 56635680 | 56645731 | 22 | 0.91 | 2.78<br>X10-07 | MT1A, 2A,<br>3, 4 |
| chr16 | Gain | 57333722 | 57335017 | 22 | 0.91 | 2.78<br>X10-07 | TRNALeu |
| chr16 | Gain | 57495650 | 57503371 | 22 | 0.91 | 2.78<br>X10-07 | POLR2C |
| chr16 | Gain | 71842906 | 71843647 | 22 | 0.91 | 2.78<br>X10-07 | AP1G1 |

| Good vs. poor prognosis CPM |  |  |  |  |  |  |  |
| --- | --- | --- | --- | --- | --- | --- | --- |
| chr3 | 6902689 | 6905031 | Loss | 12 | 1 | 2.22<br>X10-16 | GRM7 |
| chr3 | 16554466 | 16554910 | Loss | 12 | 1 | 2.22<br>X10-16 | RFTN1 |
| chr3 | 36985697 | 36986697 | Loss | 12 | 1 | 2.22<br>X10-16 | TRANK1 |
| chr3 | 43020703 | 43022351 | Loss | 12 | 1 | 2.22<br>X10-16 | FAM198A |
| chr3 | 44596684 | 44596846 | Loss | 12 | 1 | 2.22<br>X10-16 | ZKSCAN7 |
| chr3 | 46939978 | 46940439 | Loss | 12 | 1 | 2.22<br>X10-16 | PTH1R |
| chr4 | 25657365 | 25657628 | Loss | 12 | 1 | 2.22<br>X10-16 | SLC34A2 |
| chr4 | 57975793 | 57976944 | Loss | 12 | 1 | 2.22<br>X10-16 | IGFBP7 |
| chr4 | 81187125 | 81188051 | Loss | 12 | 1 | 2.22<br>X10-16 | FGF5 |
| chr4 | 81951953 | 81952024 | Loss | 12 | 1 | 2.22<br>X10-16 | BMP3 |

*Chr, chromosomes, Type CNA, Start, start position of the CNA, End, end position of the CNA, count, number of samples carrying this type of event at this location, score, proportion of samples carrying this type of event at this location, GW\_P, poisson-binomial test for overlap of event across individuals at this location given proportion chromosome affected, Gene – gene encoded by this portion of the genome.*

